# Trauma and posttraumatic stress interact with sex-specific risk loci for suicidality and converge on brain extracellular matrix biology and synaptic plasticity

**DOI:** 10.1101/2020.05.18.20105734

**Authors:** Frank R Wendt, Gita A Pathak, Daniel F Levey, Yaira Z Nuñez, Cassie Overstreet, Chelsea Tyrrell, Keyrun Adhikari, Flavio De Angelis, Daniel S Tylee, Aranyak Goswami, John H Krystal, Chadi G Abdallah, Murray B Stein, Henry R Kranzler, Joel Gelernter, Renato Polimanti

## Abstract

We performed a multivariate gene-by-environment genome-wide interaction study (GEWIS) of suicidality in 123,633 individuals using a covariance matrix based on 26 environments related to traumatic experiences, posttraumatic stress, social support, and socioeconomic status. We discovered sex-specific risk loci exhibiting interaction effects driven by PTSD, including the male-specific *rs2367967 (CWC22)*, which replicated in an independent cohort. Sex-specific cell-type transcriptome enrichment, gene-based analyses, and locus annotation point to extracellular matrix biology and synaptic plasticity as biological mediators of the effects of lifetime traumatic experiences on genetic risk for suicidality.

## Introduction

Suicide is the uniquely human act of intentionally ending one’s life. Suicide is a critical public health concern with nearly 45,000 people dying by suicide per year in the United States alone.^1^ Over 70% of individuals who die by suicide had a prior diagnosed mental disorder.^2^ Genome-wide studies have estimated the single nucleotide polymorphism (SNP)-based heritability (*h*^2^) of death by suicide to be 25-48% depending on the ancestral background of the study population ^3, 4^ Recent large-scale studies have focused on understanding the genetic risk for thoughts and behaviors leading up to the commission of acts to end ones’ life: ideation^5^ planning, and attempt.^6^ In this study we use the term suicidality to collectively refer to these thoughts and behaviors.^7^

The thoughts and behaviors leading up to suicide are interpersonally and situationally complex; and 33-98% of individuals exhibiting non-fatal suicidal behaviors have a prior psychiatric diagnosis^2^ With ~7.6% *h*^2^, a recent large-scale GWAS has detected several genome-wide significant (GWS) loci for suicidality^7^ Under the stress-diathesis model, suicide and its preceding thoughts and behaviors are the result of interaction between a stressor, environment, or state-of-being and a susceptibility to suicidal behaviors. It remains unclear which stressors are most important in influencing suicidality risk for this stress-diathesis model.^8^ Epidemiologic studies recognize that trauma and posttraumatic stress (PTS) are associated with heightened suicidal behavior severity,^9, 10, 11^ yet examination of these associations from a genetic perspective is limited.

Gene-by-environment (GxE) interaction studies historically rely on *a priori* hypotheses regarding SNP or gene effects on a phenotype given specific environmental conditions.^12, 13^ Large-scale genetic studies fail to support historical candidate genes and GxE interactions.^14^ Alternatively, genome-wide gene-by-environment interaction studies (GEWIS) are suitable for detection of risk loci across the genome, while assessing interactions with multiple, potentially correlated, environmental factors.^15^

We performed a multivariate GEWIS of a derived ordinal suicidality phenotype in 123,633 participants^7^ (55.9% female) from the UK Biobank (UKB) using a linear mixed model to investigate a covariance matrix based on 26 environments related to traumatic experiences, PTS, social support, and socioeconomic status. From GEWIS of the full UKB cohort and the sex-stratified analyses, we discovered five suicidality risk loci with interaction effects primarily mediated by PTS and PTSD. We further characterized these risk loci and identified sex-specific effects with convergent evidence of extracellular matrix biology and synaptic plasticity as biological mediators of the relationship between genetic risk for suicidality and PTS.

## Results

### Cohort and phenotype summary

After sample and variant quality control we analyzed 7,284,651 SNPs for suicidality^7^ GxE effects in 123,633 participants (55.9% female) of European ancestry from the UKB. Of these, 84,196 (52.7% female) were coded as “no suicidality” controls (rank=0), 21,176 (60.7% female) as “thought life not worth living” (rank=1), 13,078 (63.6% female) as “contemplated self-harm or suicide” (rank=2), 2,487 (70.7% female) as “deliberate self-harm” (rank=3), and 2,696 (68.1% female) as “attempted suicide” (rank=4) (Figure 1).^7^

**Figure 1.**
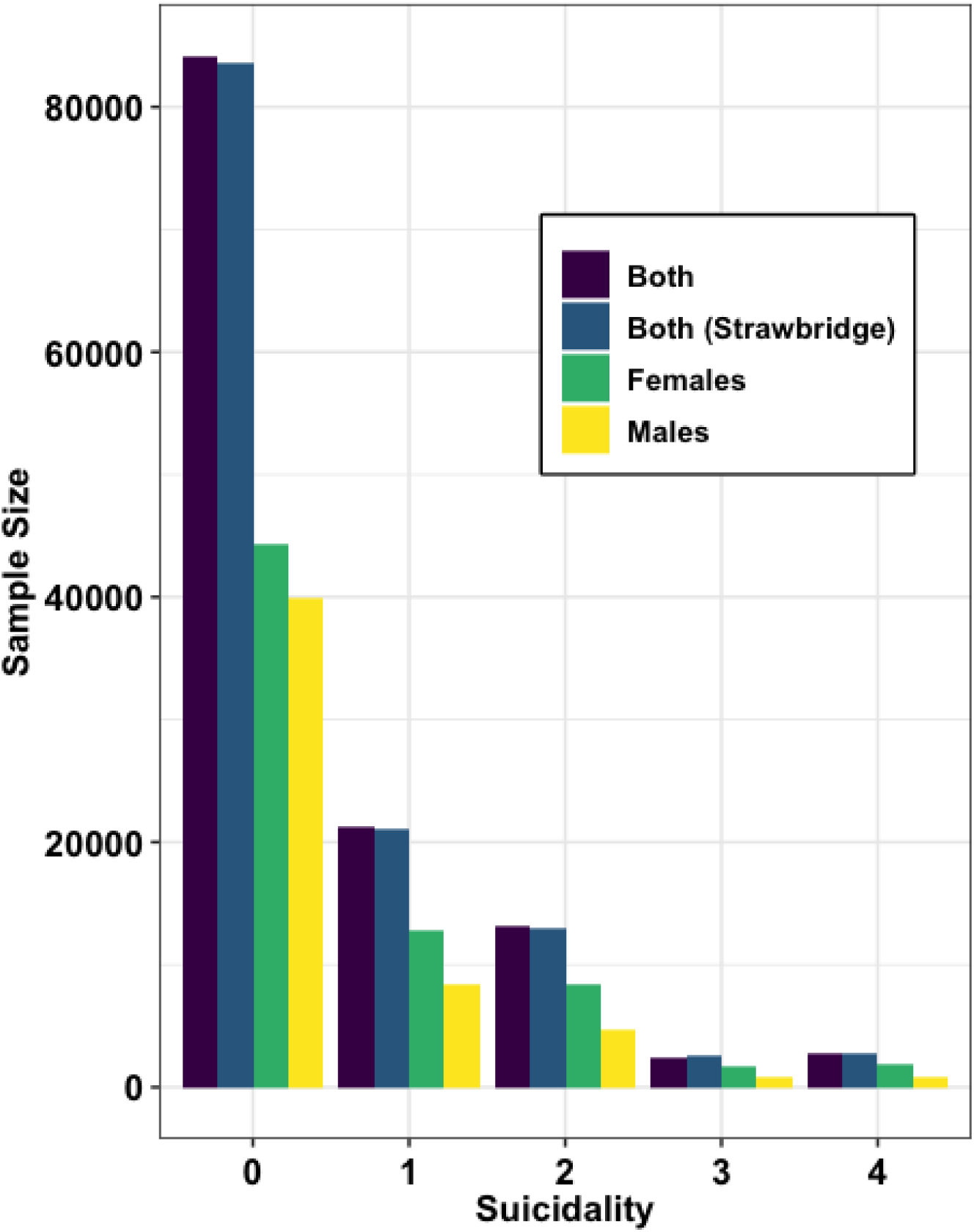
Sample sizes used in this GEWIS of suicidality by sex and in the full cohort and those used in the GWAS by Strawbridge, *et al*.^7^ Suicidality is coded as follows: 0 = no suicidality controls, 1 = “thought life not worth living,” 2 = “contemplated self-harm or suicide,” 3 = “deliberate self-harm,” and 4 = “attempted suicide.”

Missing data from the UKB for all 26 environments were imputed using Multivariate Imputation via Chained Equations (MICE)^16, 17^ using ten iterations of 5 multiple imputations. For 24 out of 26 environments (described in Tables S1 and S2), less than 10% of participants required imputation. Two environments, *“felt irritable or had angry outbursts in past month”* and *“felt distant from other people in past month,”* had missing information for 68,972 and 68,699 individuals, respectively. After imputation of all 26 environments, there was a relative decrease in standard error estimates indicating that the data were reliably imputed (Figures S1 and S2, Tables S1 and S2).^16, 17^

All environments were tested for phenotypic and genetic correlation with suicidality (Figures 2 and S3). After multiple testing correction (*FDR*<5%), all environments were associated with suicidality in males, females, and the full cohort in generalized linear models covaried for age, sex, genotype batch, and ten principal components (PCs) of ancestry. GWAS summary statistics for each environment were accessed via the UKB (see http://www.nealelab.is/uk-biobank) and formatted with Linkage Disequilibrium Score Regression (LDSC) using the 1000 Genomes Project European populations as a linkage disequilibrium reference panel.^18^ GWAS summary statistics for suicidality were retrieved from Strawbridge, *et al*.^7^ All environments and the ordinal suicidality phenotype exhibited significant heritability (*h*^2^) with *h*^2^ z-scores ≥ 4, indicating sufficient power for genetic correlation. After multiple testing correction *(FDR<5%)*, suicidality was genetically correlated with all but four environments: UKB Field ID 738: *average total household income before tax*, UKB Field ID20525: *able to pay rent/mortgage as an adult*, UKB Field ID 20527: *been involved in combat or exposed to war-zone*, and UKB Field ID 20491: *someone to take you to doctor when needed as a child* (Figure S3).

**Figure 2.**
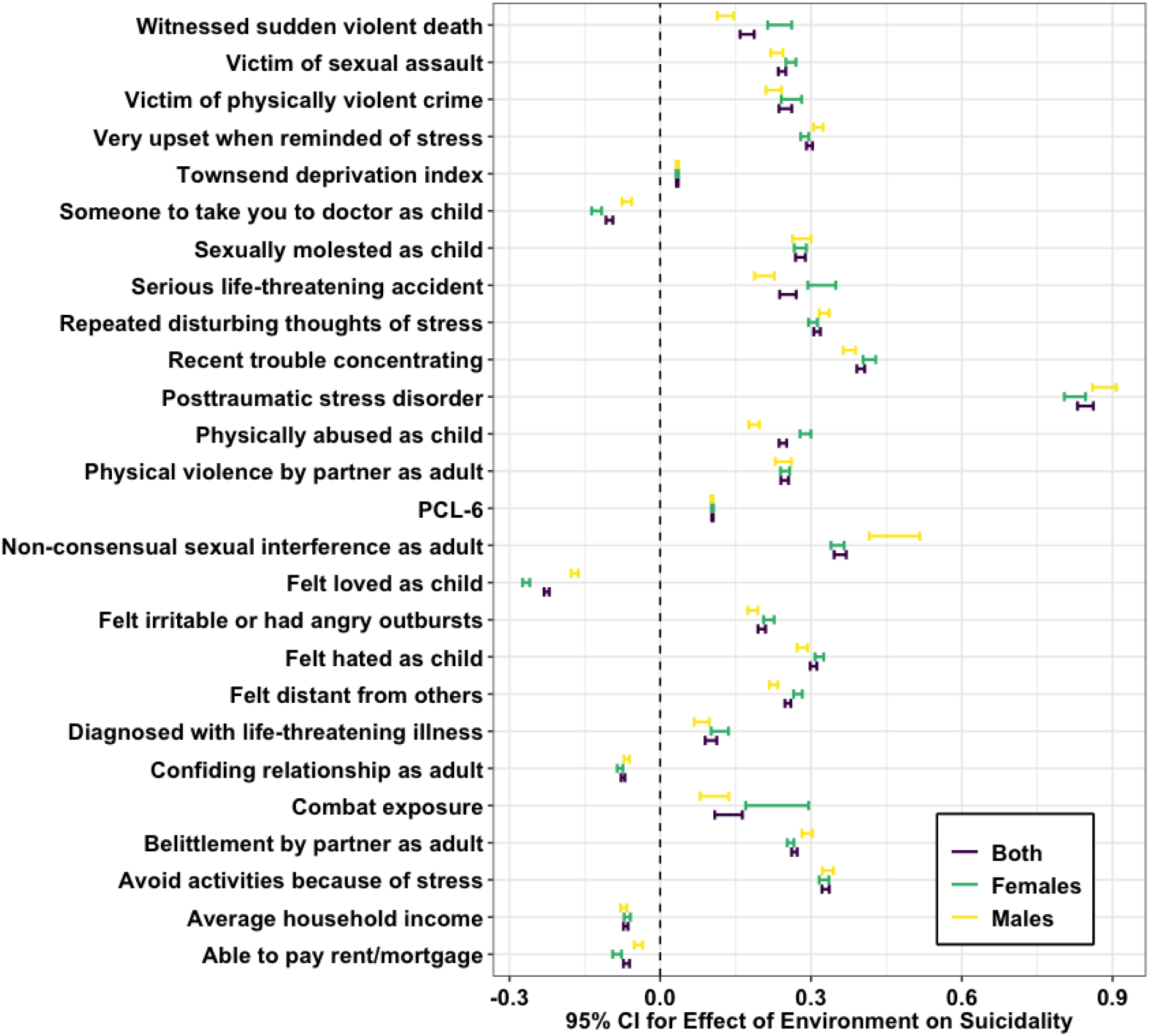
Association between suicidality and 26 environments related to traumatic experiences, responses to trauma, posttraumatic stress, social support, and socioeconomic status. Shown are 95% confidence intervals surrounding point estimates from a generalized linear model of suicidality adjusting for age, sex, genotyping batch, and ten principal components of ancestry.

### Suicidality Risk Locus Discovery using multivariable GEWIS

Multivariate GEWIS in StructLMM^15^ is capable of evaluating GxE with large a covariance matrix so we included all 26 imputed environments in our discovery GEWIS. In the main discovery GEWIS of 126,633 individuals we detected one risk locus in association and interaction tests whose lead SNP was *rs12589041* (*p_association_* (*p_a_*)=2.43×10^-8^, *p_interaction_* (*p_int_*)=2.04×10^-9^; Table 1 & Figure S4). This SNP maps to the region of the genome coding for the long non-coding RNA *RP6-65G23.1*. In sex-stratified GEWIS we discovered 1 significant risk locus for suicidality in females (*rs118118557 p_a_*=2.37×10^-8^, *p_int_*=1.14×10^-7^; Figure S5) and three LD independent risk loci for suicidality in males (*rs2367967 p_a_*=2.47×10^-9^, *p_int_*=5.91×10^-10^, *rs72619337 p_a_*=1.08×10^-8^, *p_int_*=1.63×10^-9^; and *rs6854286 p_a_*=1.98×10^-8^, *p_int_*=9.41×10^-9^; Figure S6). The *rs118118557* risk locus detected in females positionally mapped to the transcription start site of *CHST14*, which encodes carbohydrate sulfotransferase 14, an essential enzyme for extracellular matrix production in the hippocampus and the proliferation and neurogenesis of neural stem cells.^19^ The *rs2367967* locus discovered in males positionally mapped to the *CWC22* locus, which encodes pre-mRNA-splicing factor CWC22 homolog.^20, 21, 22^ CWC22 is a spliceosomal protein responsible for controlling neurite outgrowth in developing neurons.^23^

**Table 1.** Lead SNP information for five risk loci, including tested alleles (A1) and their frequencies (AF) in UKB, detected in a GEWIS of suicidality in the UKB.

| RSID | Sex | Chr | Position | $P_{\text{association}}$ | $P_{\text{interaction}}$ | AF (A1) | Nearest Gene |
| --- | --- | --- | --- | --- | --- | --- | --- |
| rs12589041 | Both | 14 | 71172013 | $2.43 \times 10^{-8}$ | $2.04 \times 10^{-9}$ | 0.25 (G) | RP6-65G23.1 |
| rs118118557 | Female | 15 | 40762879 | $2.37 \times 10^{-8}$ | $1.14 \times 10^{-7}$ | 0.01 (G) | CHST14 |
| rs2367967 | Male | 2 | 180948951 | $2.47 \times 10^{-9}$ | $5.91 \times 10^{-10}$ | 0.32 (G) | CWC22 |
| rs6854286 | Male | 4 | 12652325 | $1.98 \times 10^{-8}$ | $9.41 \times 10^{-8}$ | 0.37 (C) | RP11-145E17.2 |
| rs72619337 | Male | 9 | 137175269 | $1.08 \times 10^{-8}$ | $1.63 \times 10^{-9}$ | 0.03 (G) | RP11-352E6.1 |

### Comparing GWAS vs. multivariable GEWIS of suicidality

Strawbridge, *et al*.^7^ conducted a GWAS not accounting for GxE effects and detected three GWS loci associated with suicidality. In our multivariable GEWIS interaction tests, these three loci showed nominally significant GxE interactions with the covariance matrix tested (*rs62535711 p_int_*=6.76×10^-5^; *rs598046 p_int_*=2.00×10^-6^; *rs7989250 p_int_*=0.023). In the multivariable GEWIS association tests, *rs598046* also was associated with suicidality (*p_a_*=0.020) (Table S3). Because of the GxE effects at these loci,^7^ we included them in the subsequent GxE characterization analyses.

The effects of loci identified in our multivariate GEWIS were not detected in Strawbridge, *et al*,^7^ possibly because environmental heterogeneity was not accounted for in that GWAS.

### Locus Replication in Yale-Penn using multivariable GEWIS

In the independent Yale-Penn replication sample (*N*=3,099, 37.5% female)^24, 25, 26, 27, 28^ using environments encompassing comparable domains as UKB (see Online Methods *Replication)* we performed association and interaction tests with StructLMM^15^ for all SNPs in each genomic risk locus discovered with GEWIS. Although the Yale-Penn sample is much smaller than the UKB discovery sample, the male-specific genomic risk locus mapping to *CWC22* replicated (*N*=1,936 males; *rs2367967*, Yale-Penn *p_a_*=0.019, *p_int_*=0.099). In the same genomic region, we observed an additional signal partially independent from *rs2367967*: *rs17781395* (chr2:180927110; *D*’=1.0, *R*^2^=0.177) was associated with suicidality risk with non-significant evidence of GxE interaction (*p_a_*=8.99×10^-7^, *p_int_*=0.1). The *rs12589041* proxy SNP *rs7145623* (*D*’=1.0, *R*^2^=0.995) was not replicated in the Yale-Penn cohort (*p_a_*=0.611, *p_int_*=0.376). The Yale-Penn cohort did not contain genotype information for *rs118118557, rs6854286, rs72619337*, or a suitable LD proxy variant (*R*^2^≥0.5) (Table S5-9).

### Major Environmental Mediators

All GEWIS-discovered loci showed compelling evidence for interaction with *posttraumatic stress disorder (PTSD)* and/or a quantitative *PTSD* risk score (the PTSD Checklist 6-tem subset, *PCL-6)* (Figure 3 and S7): GxE for *rs12589041* in both sexes had log(Bayes factor (*BF*))=9.97 for *PTSD* and log(*BF*)=9.59 for *PCL-6;* GxE for *rs118118557* in females had log(*BF*)=8.82 for *PTSD* and log(*BF*)=8.86 for *PCL-6;* in males, GxE at *rs2367967* had log(*BF*)=5.27 for *PTSD* and log(*BF*)=6.56 for *PCL-6*, GxE at *rs6854286* had log(*BF*)=10.41 for *PTSD* and log(*BF*)=5.31 for PCL-6, and GxE at *rs72619337* had log(*BF*)=8.04 for *PTSD* and log(*BF*)=10.57 for *PCL-6*.

**Figure 3.**
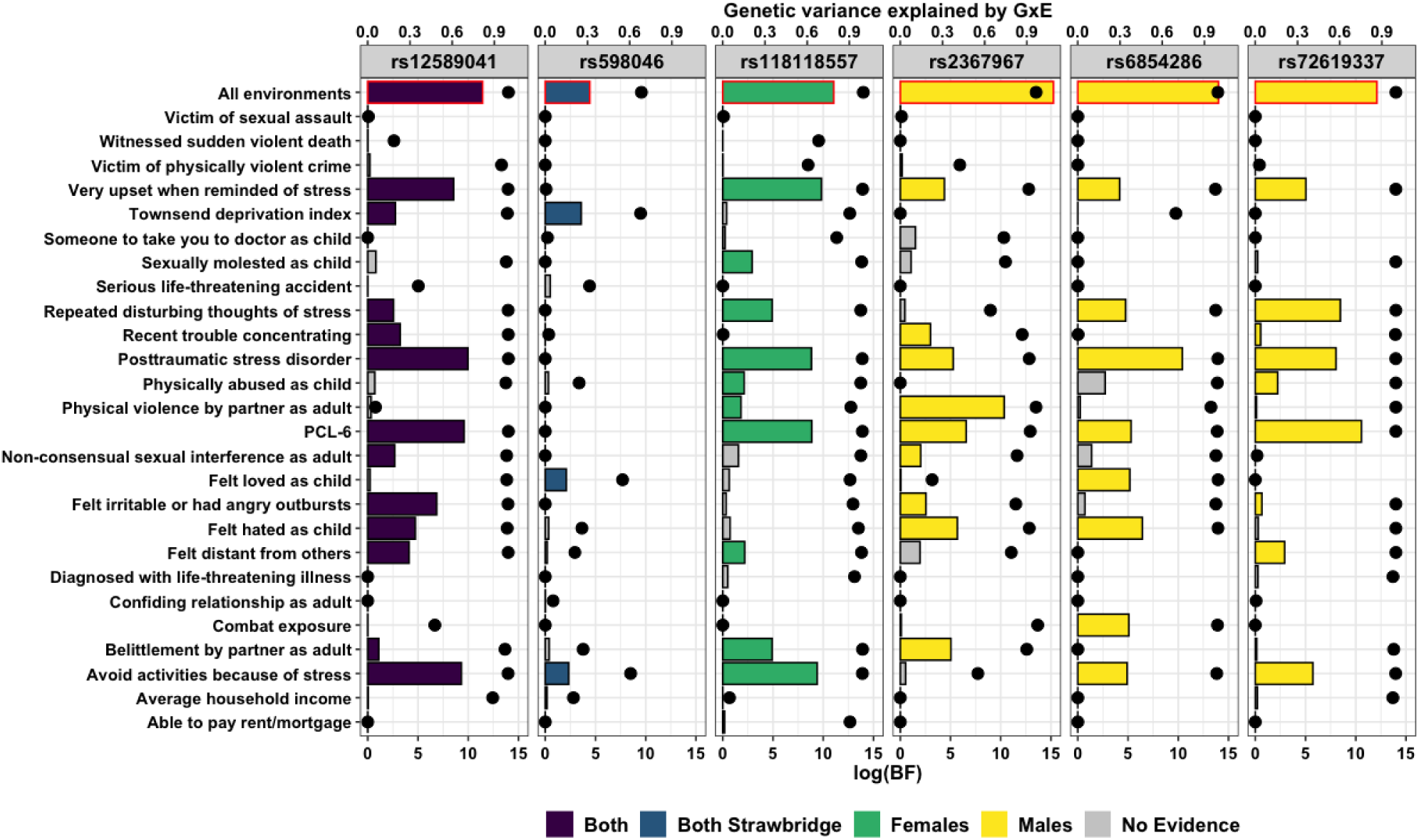
Individual relevance of the environment included in the covariance matrix used to detect gene-by-environment (GxE) interactions with suicidality. Each bar represents the log marginal likelihood (log(Bayes factor(*BF*))) of models for the indicated environment relative to models lacking that environment. Black data points indicate the genetic variance explained by GxE at each locus for the indicated environment. Bars outlined in red highlight the full GxE model at each locus; colored bars indicate environments with positive evidence for their contribution as part of a causal set of environments underlying the detected GxE (Figure S7).

Consistent with Moore, *et al*.,^15^ we identified many environments driving the detected GxE interactions (Figure 3) but we also observed distinct GxE architectures per SNP after greedy backwards elimination of most probably causal interacting environments included in the covariance matrix. All probably causal environment sets contained PCL-6 and/or PTSD. In females, two additional environments were deemed possibly causal for GxE at *rs118118557*: UKB Field ID 20498 *felt very upset when reminded of stressful experience in past month* log(*BF*)=9.82 and UKB Field ID 20495: *avoided activities or situations because of previous stressful experience in past month* log (BF)=9.39. Implicated causal environments in males include UKB Field ID 20523: *physical violence by partner or ex-partner as an adult* (log(*BF*)=10.34 at *rs2367967)*, UKB Field ID 20487: *felt hated by family member as a child* (log(*BF*)=6.42 at *rs6854286)*, and UKB Field ID 20497: *repeated distributing thoughts of stressful experience in past month (i.e*., re-experiencing; log(*BF*)=8.47 at *rs72619337)*.

One suicidality risk locus from Strawbridge, *et al*.^7^ showed evidence of GxE interaction with the environments evaluated in this study *(rs598046* full model log(*BF*)=4.43; *rs62535711* full model log(*BF*)=0; and *rs7989250* full model log(*BF*)=0). The GxE at *rs598046 (CNTN5* intronic variant) was driven primarily by UKB Field ID 189: *Townsend deprivation index at recruitment* (log(*BF*)=3.58, Figure S7).

### Genetic Variance Explained by GxE

Next, we evaluated the genetic variance at each implicated locus explained by GxE (rho; *ρ*) accounting for the full environment model (*N*=26) and each environment individually (Figure 3). The full GEWIS explained nearly all of the genetic variance at all five GEWIS discovered SNPs: *rs118118557 ρ*=99.9%, *rs12589041 ρ*=99.9%, *rs2367967 ρ*=96.6%, rs6854286 *ρ*=99.6%, and rs72619337 *ρ*=99.9%. As expected, the genetic variance explained at loci detected by Strawbridge, *et al*.^7^ were much lower than those detected by GEWIS because these variants were originally detected in a GWAS that did not account for GxE effects (*rs598046 ρ*=68.4%; *rs62535711 ρ*=6.85%; *rs7989250 ρ*=2.10%).

### Effect Size Predictions

Effect sizes were predicted in a test sample comprised of 50% of the total sample and calculated with all 26 environments from the original model using the best linear unbiased predictor (BLUP).^15^ All five suicidality risk loci discovered by GEWIS exhibit qualitative interactive effects in which the effect of each SNP on suicidality switches direction depending on an individual’s environmental profile (Figure 4). Two loci exhibited positive persistent genetic effects on suicidality: *rs118118557* in females (*β*=1.39×10^-3^) and *rs72619337* in males (*β*=0.011) and three loci exhibited negative persistent genetic effects: *rs12589041* in the full cohort (*β*=-1.49×10^-3^), *rs2367967* in males (*β*=-4.62×10^-3^), and rs6854286 in males (*β*=-9.35×10^-3^).

**Figure 4.**
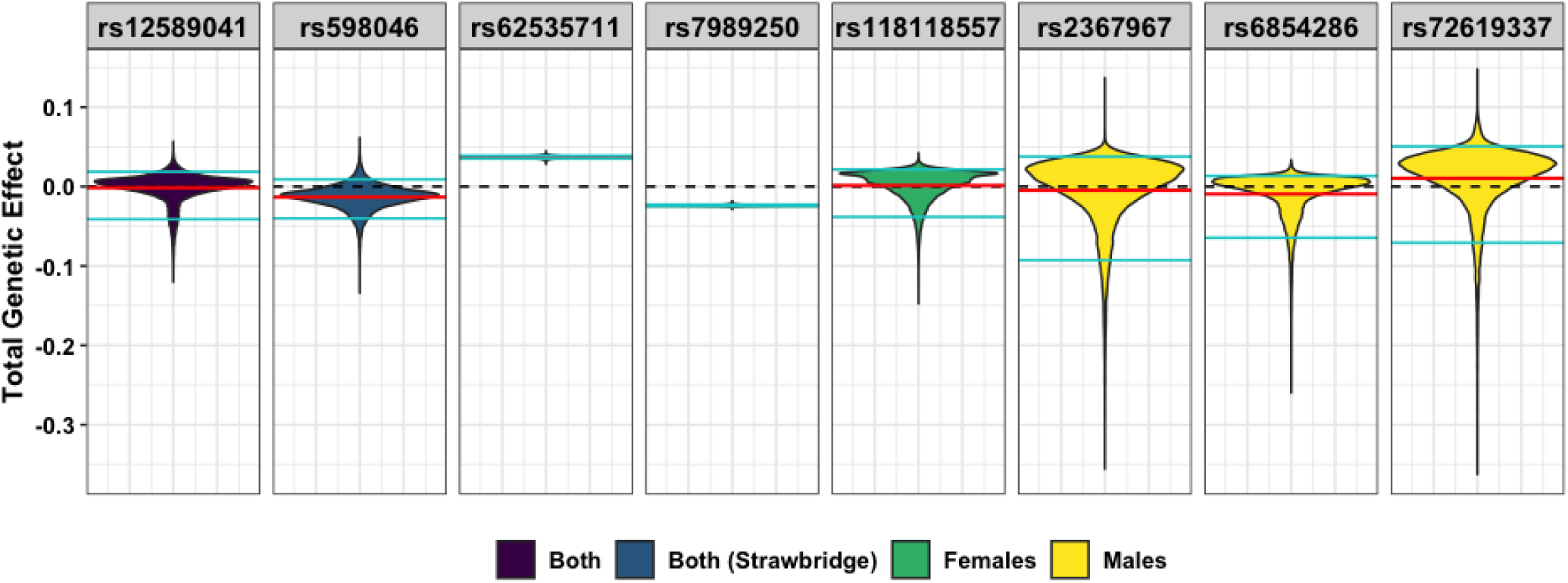
Predicted genetic effects of GEWIS loci for suicidality. Violin plots of the distribution of estimated allelic effect sizes at each locus in 61,816 unrelated individuals (“Both”), 27,256 unrelated males, and 34,560 unrelated females of European ancestry given full models of traumatic events and posttraumatic stress environments. Persistent genetic effect estimates are shown in solid red lines; zero effect is shown in black dashed lines; cyan lines indicate the top and bottom 5% of each distribution.

Effect sizes for the loci that Strawbridge, *et al*.^7^ detected by GWAS should show effects on suicidality unbiased by GxE effects. GEWIS persistent genetic effects for *rs62535711* (*β_GEWIS_*^=^0.037, *β_strawbridge_=*0.105, *p_diff_*=1.70×10 ^4^) and *rs7989250 (β_GEWIS_=-* 0.024, *β_Strawbridge_*=-0.052, *p_diff_*=0.002) were consistent, yet significantly smaller, effect estimates compared to GWAS. The locus *rs598046 (CNTN5* intronic variant) demonstrated qualitative interactive effects given trauma and PTS environments with a persistent genetic effect significantly different than, and in the opposite direction from, that reported by Strawbridge, *et al*.^7^ (*β_GEWIS_*=-0.013, *β_strawbridge_*=0.053, *p_diff_*=1.84×10^-13^; Figure 4).

### Brain Tissue Measurements

We investigated potential neurobiological underpinnings of the detected GxE relationships by testing each GEWIS locus for GxE effects in brain phenotypes from UKB (872 brain-imaging phenotypes; *N≤*39,755). After multiple testing correction for the number of tests performed per sex (*FDR*<5%), we detected 285 brain-imaging correlates of suicidality in the full cohort: 189 in females, and 33 in males (Table S10-12). These brain-imaging phenotypes were each tested individually for GxE interaction with each suicidality risk locus including age, genotyping batch, and ten ancestry PCs as covariates. For tests of risk loci in the full cohort, sex also was included as a covariate. We detected nominally significant GxE-adjusted associations at *rs12589041* in the full sample putatively implicating various regions of the brain involved in memory *(e.g*., volume of hippocampus *p_a_*=0.037), emotions *(e.g*., volume of grey matter in amygdala *p_a_*=0.040), and goal-oriented behaviors *(e.g*., weighted-mean fractional anisotropy in tract inferior fronto-occipital fasciculus *p_a_*=0.049) (Tables S13-15).

### Gene-based Association Tests

Gene-based tests were performed with MAGMA (Multi-marker Analysis of GenoMic Annotation)^29^ in FUMA (Functional Mapping and Annotation of Genome-Wide Association Studies)^30, 31^ by combining single-variant signals within genic regions accounting for LD structure. In the full sample (UKB *N*=123,633), three genes were associated with suicidality in association and interaction tests after multiple testing correct (*FDR*<5%; Figure 5 and Table S16): *CNTN5* (contactin-5, *z_a_*=5.86, *p_a_*=2.38×10^-9^, *z_int_*=2.98, *p_int_*=1.51×10^-3^), *PSMD14* (26S proteasome non-ATPase regulatory subunit 14, *z_a_*=5.07, *p_a_*=2.04×10^-7^, *z_int_*=4.32, *p_int_*=7.76×10^-6^), and *HEPACAM* (hepatocyte cell adhesion molecule, *z_a_*=4.57, *p_a_*=2.43×10-^6^, *z_int_*=4.96, *p_int_*=3.82×10^-7^). *CNTN5* was previously implicated in suicidality^7^ and was discovered herein to harbor a suicidality risk SNP that interacted with traumatic events and posttraumatic stress.

**Figure 5.**
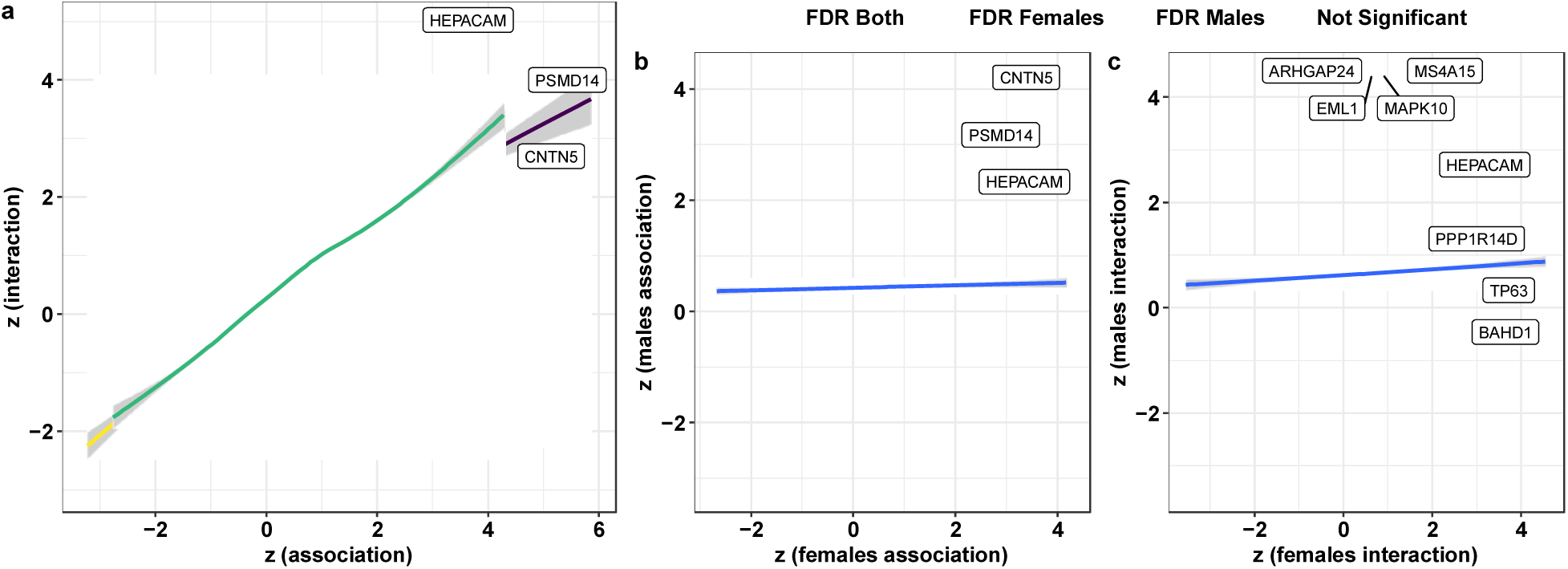
Gene-based GEWIS results. (a) Comparison of association and interaction gene-based test statistics. Labeled genes were significantly associated with suicidality in association and interaction GEWIS. (b,c) Comparison of gene-based associations between males and females. In b, no gene survived multiple testing correction in either analysis so the genes from (a) are labeled. In c, genes differentially associated with suicidality between males and females are labeled. Linear regression between male and female association and interaction tests are shown in blue lines.

Sex-stratified analyses recapitulate findings from the full sample (Figure 5 and Table S16), identifying non-overlapping significantly associated genes in interaction tests. Genes significantly associated with suicidality after FDR correction in males and females were analyzed for gene ontology enrichment in ShinyGO v0.61 (Table S17)^32^. The female-associated genes exhibited enrichment of chromatin remodeling (GO:0006338 *FDR q*=0.011) while male-associated genes exhibited enrichment of diabetes (type II diabetes mellitus *FDR q*=0.029) and loss of previously present mental ability (particular in adults; Human Phenotype Ontology mental deterioration (HP:0001268) *FDR q*=0.034).^33^

### Functional Annotation and Regulatory Effects

Using genome-wide information from the association and interaction GEWIS analyses, we tested for gene-set, cell-type transcriptomic profile, and tissue-transcriptomic profile enrichments as well as expression quantitative trait locus (eQTL) and chromatin interactions in males, females, and the full sample. After multiple testing correction (*FDR*<0.05), the GEWIS association test in males was enriched for two gene-sets: mature B-cell differentiation (*FDR q*=0.033)^34^ and response to testosterone (*FDR q*=0.049) (Table S18). After multiple testing correction for the number of within-data set cell types (*FDR*<0.05), we detected three middle temporal gyrus (MTG) cell-type transcriptomic profiles enriched in the GEWIS of suicidality in males (Table S19): (1) *SCUBE3-* and *PVALB*-expressing inhibitory neurons from MTG layers 2-5 (association tests: *β*=0.028, *FDR q*=0.028; interaction tests: *β*=0.018, *FDR q*=0.049), (2) *OPRM1-*expressing neurons from MTG layers 1-4 (*β*=0.030, *FDR q*=0.049), and (3) *SPAG17-* expressing inhibitory neurons from MTG layers 2-4 (*β*=0.029, *FDR q*=0.049).

In the full UKB sample, the genomic risk locus containing *rs12589041* was a significant PsychENCODE eQTL for *TTC9* (tetratricopeptide repeat domain 9; *FDR q*=0.026) but did not exhibit brain-associated chromatin interactions (Figure S8 and Table S20). In females, the genomic risk locus containing *rs118118557* exhibited several significant chromatin interactions in both fetal and adult cortex, the most significant of which involved the genomic region containing *CCDC32* (fetal cortex *FDR q*=2.61×10^-35^; adult cortex *FDR q*=1.25×10^-46^; Figure S9 and Table S21). In males, two SNPs in the genomic risk locus containing *rs2367967* were significant PsychENCODE eQTLs for *UBE2E3 (FDR q*=0.044 and 0.046; Figure S10 and Table S22). This same genomic risk locus exhibited significant chromatin interactions with the region of chromosome 2 containing *NEUROD1* and *CERKL* (dorsolateral prefrontal cortex cells *FDR q*=6.75×10^-10^), *ITGA4* (neural progenitor cells *FDR q*=1.27×10^-8^), and *ZFN385B* (neural progenitor cells *FDR q*=4.07×10^-19^; Figure S10 and Table S23). In males, the genomic risk locus containing *rs6854286* exhibited significant chromatin interaction with the chromosome 2 region encoding *HS3ST1* (neural progenitor cells *FDR q*=1.18×10^-7^; Figure S11 and Table S23).

## Discussion

We conducted a comprehensive analysis of suicidality describing (1) five GEWIS risk loci with interaction and association effects with specific environments including sex-specific GxE effects, (2) environments considered possibly causal for the GxE interactions at these loci, (3) replication of locus-suicidality associations in an independent dataset, (4) description of GxE effects that may have confounded prior large-scale studies of suicidality, and (5) annotation of genome-wide suicidality genetic architecture uncovering relevant neuronal cell types and brain-circuitry. In aggregate, these data identify a pinpoint potential molecular target, namely the extracellular matrix (ECM), for suicidality that interacts with traumatic experiences and posttraumatic stress.

Of the five suicidality risk loci that we discovered by GEWIS, two mapped to genic regions: *rs118118557* mapped to *CHST14* and *rs2367967* mapped to CWC22. *CHST14* encodes carbohydrate sulfotransferase 14 and is responsible for sulfation of *N*-acetylgalactosine residues of dermatan sulfate in the ECM^35^ ECM in the brain plays a major role in the storage of information through learning and thus, ECM maintenance, regulation, and composition are reportedly highly relevant for disorders related to, or exacerbated by, prior experiences including those traumatic events studied herein^36, 37^ Describing the GxE at the *CHST14* SNP *rs118118557* through greedy backwards elimination *(i.e*., stepwise removal of environments with highest log likelihood relative to a hypothesis of no GxE effects) of environments demonstrated an interaction with physically and sexually violent experiences in childhood and adulthood.^15^ This relationship suggests that synaptic plasticity mechanisms are possible targets for treating suicidality as well as psychiatric disorders with a strong influence from memories of prior experiences^38^ such as PTSD,^39^ addiction,^40^ major depressive disorder,^39^ and schizophrenia.^41^ Possible synaptic plasticity associated targets include decentering: the capacity to observe items arising in the mind *(e.g*., thoughts, feelings, memories, or bodily sensations) with physiological distance, heightened self-awareness, and perspective thinking as competencies that may reduce the risk of suicidality.^42^

*CWC22* encodes the “CWC22 spliceosome associated protein homolog” protein, which is the only protein-coding component of the large (127kb) Rd2 segmental duplication. *CWC22* is one of the protein-coding regions of the genome that is most often duplicated^43^ Large segmental duplications are often enriched for environmentally responsive genes^44^ and *CWC22* is no exception. *CWC22* and the Rd2 segmental duplication have previously been implicated in brain microstructural changes in response to exercise^45^ and social engagement^46^ both of which exhibit memory-inducing effects on the ECM.^47, 48^ Environmental enrichment (EE) describes exposure to situations that facilitate enhanced sensory, cognitive, and motor stimulation and is known to affect synaptic plasticity in humans^46^ but is most often implemented in rodents^49^ We hypothesize that the association between *CWC22* and suicidality suggests that such a noninvasive, non-pharmacological, and personalizable treatment strategy such as EE may effectively mitigate the effects of genetic liability to suicidality.

Gene-based association testing is more powerful than per-SNP tests because they combine multi-locus signals into a single genic region, reducing the burden of multiple testing correction.^29^ Using this approach we discovered three genes associated with suicidality: *HEPACAM* (hepatocyte cell adhesion molecule), *PSMD14* (26S proteasome non-ATPase regulatory subunit 14), and *CNTN5* (contactin-5). *HEPACAM* encodes GlialCAM, which is responsible for promoting interactions between ECM components and glia^50^ *HEPACAM* harbors at least 20 SNPs associated with megalencephalic leukoencephalopathy with subcortical cysts, a disorder characterized by an enlarged brain and abnormally myelinated white matter^51, 52^ The ubiquitin-proteasome system, of which the *PSMD14* gene product is a component, plays a critical role in the expression of ECM-associated genes^53^ Most notable is *CNTN5*, previously detected by a SNP-based GWAS of suicidality also performed in the UKB – a finding that is consistent, but that made use of some of the same information included herein. Its product, contactin-5 is part of the glycosylphosphatidylinositol-bound protein family that has often been implicated in autism spectrum disorder.^54^ Contactin-5 is secreted into the ECM, where it facilitates the formation and growth of neuronal projections^55^ Strawbridge, *et al*.,^7^ described the *rs598046* risk locus, mapped to *CNTN5*, as conferring increased risk for suicidality (*β*=0.053).^7^ However, using GEWIS, we observed significant interaction effects at *rs598046* whereby across the range of Townsend deprivation index scores, *rs598046* conferred increased *and* decreased suicidality risk. The confounding nature of socioeconomic status has been observed extensively in GWAS of psychiatric disorders including PTS and PTSD^56, 57, 58, 59^ and is therefore unsurprising to observe similar effects by GEWIS. This finding highlights the importance of GEWIS and the GxE characterization of GWAS risk loci to adequately evaluate the effects of environmental heterogeneity, especially via the effects of socioeconomic status.

Detected via functional annotation of the suicidality GEWIS in males only, we recapitulate two cell types previously implicated in suicidal behaviors: *SPAG17-*expressing and *OPRM1*-expressing inhibitory neurons from the middle temporal gyrus^60^ *SPAG17* (sperm-associated antigen 17) was previously identified by whole exome sequencing of suicide deaths as a locus enriched for rare variants.^61^ *OPRM1* encodes the mu-opioid receptor 1 and has garnered considerable attention in candidate gene and large-scale genetic studies of substance use and abuse,^62^ schizophrenia,^63^ Tourette syndrome,^64^ suicidal ideation,^65^ and suicidal behavior.^66^ We also detected inhibitory neurons from layers 2-5 of the middle temporal gyrus expressing both *SCUBE3* and *PVALB*.^60^ *PVALB* encodes parvalbumin, a calcium-binding protein that supports neuroplasticity and other functional properties of fast-spiking interneurons in cortical and subcortical regions, influencing stress response in animals.^67,68,69^ In humans, pathology in parvalbumin neurons was implicated in schizophrenia,^70^ depression,^71^ Gilles de la Tourette^72^ syndrome, and other psychiatric disorders. These findings highlight genetic and biological overlap between suicidality and key genetic targets for other psychiatric disorders, especially those exacerbated by adverse prior experiences, and suggests that there are pervasive environmental interaction effects underlying genetic risk for the trait.

Several regulatory interactions were detected that further implicate the ECM and synaptic plasticity in the GxE effect of posttraumatic stress and genetic risk for suicidality. Reduced *TTC9* (tetratricopeptide repeat domain 9) expression is associated with higher anxiety-like behaviors^73, 74, 75^ and regulatory pathways implicated in adult neuroplasticity and serotonin signaling. CCD32 (coiled-coil domain containing 32) has been linked to activity-dependent synaptic plasticity in mature olfactory neurons.^76^ NEUROD1 (neuronal differentiation 1) is a highly potent transcription factor capable of controlling neuronal fate under some circumstances. For example, NEUROD1 alone was capable of converting reactive glia into functional neurons (*in vivo*).^77^ NEUROD1 activates neuronal development genes though direct replacement of repressive methyl groups with activating acetyl signatures. Transient activity of NEUROD1 during development was sufficient for long-term epigenetic memory^78^ of transcriptional activity. CERKL (ceramide kinase like) has been previously implicated in genetic studies of anhedonia and perceptual aberration.^79, 80^ *HS3ST1* encodes heparan sulfate 3-O-sulfotransferase 1, which is responsible for sulfation of heparan in the ECM. Heparan sulfate has been previously implicated in ECM contributions to schizophrenia and bipolar disorder.^81^ These findings not only reinforce suicide as a psychiatric condition but also demonstrate substantial overlap between regulatory effects across disorders with larger available sample sizes and well-characterized biology.

Convergent evidence that ECM biology underlies suicidality is intriguing given recent evidence that ECM is involved in memory retention and synaptic plasticity following traumatic experiences^36^ Indeed, posttraumatic stress and subsequent PTSD were identified as key environments in all of our GEWIS-discovered loci. Rather than identifying a single environment or state-of-being that mediates GxE effects at the loci discovered herein, we uncover patterns or profiles of highly correlated (phenotypically and genetically) environments that contributed to a locus’ effect on suicidality. Many of the environments that we studied are malleable and modifiable, lending additional evidence that an EE approach could mitigate suicidality risk. For example, attachment style *(e.g*., an individual’s perception and expectations of the availability and/or responsiveness of meaningful others during times of stress^82^) is a potentially modifiable risk factor for PTS, PTSD, and suicidality. A secure attachment style is clinically a protective factor for PTSD and related symptoms^83^ but also protects against PTSD polygenic risk (unpublished data). Possible future directions for this work include the investigation of causal relationships between attachment style and social support, posttraumatic stress, and longitudinal reports of suicidal behaviors.

Our study detected novel biological underpinnings of suicidality but has some limitations. First, replication of GEWIS was limited to independent samples with comparable environments. Indeed, our replication with the Yale-Penn cohort lacked 1-to-1 matched environments; however, we successfully included variables from Yale-Penn that capture the same domains of environment or state-of-being: traumatic experiences, posttraumatic stress, social support, and socioeconomic status. The highly correlated nature of these environments suggests that GxE tests in Yale-Penn should be well suited to detect comparable association and interaction effects.

We identified interactions between suicidality risk variants and potentially modifiable environments that point to the ECM as a possible source of suicidality risk in relation to learned behavior following traumatic experiences across the lifespan. There were shared genetic effects between the identified loci and brain circuitry associated with planning behaviors and cell types that contribute to synaptic plasticity and axonal innervation. Characterization of the molecular basis for the effects of traumatic experience and posttraumatic stress on the risk of suicidal behaviors may help to identify novel targets for which more effective treatments can be developed for use in high-risk populations.

## Methods

### Discovery Cohort – UK Biobank Description and Participant Inclusion

The UK Biobank (UKB) is a large population cohort comprised of nearly 502,000 participants. UKB participants range in age from 37-73 years at time of recruitment and represent a general sampling of the UK population with no enrichment for specific disorders. Assessments included physical health, anthropometric measurements, and sociodemographic characteristics. By August 2017, 157,366 participants in the UK Biobank completed an ancillary online mental health questionnaire^84^ covering topics of self-reported mental health and well-being. From this mental health questionnaire, we defined a suicidality phenotype for 123,633 individuals described below and following the sample inclusion pipeline of Strawbridge, *et al*.^7^ plus selection of traumatic event and trauma response endorsements.

We excluded all UKB participants who were not of self-reported white British ethnicity and those who were outliers with respect to heterozygosity and missingness. From each related pair of individuals (kinship coefficient > 0.042; second cousins) we selectively removed participants endorsing the less severe suicidality symptom group to prioritize retention of persons with more severe suicidality rating. The total sample included 123,633 UKB participants. Of these, 84,196 were coded as “no suicidality” controls, 21,176 as “thought life not worth living,” 13,078 as “contemplated self-harm or suicide,” 2,487 as “deliberate self-harm,” and 2,696 as “attempted suicide.”

### UKB Suicidality

Ordinal suicidality categories were determined based on the sample inclusion criteria from Strawbridge, *et al*.^7^ Briefly, suicidality groups were based on four questions from the UKB online mental health questionnaire describing non-overlapping severity of suicidality. Based on individual responses to these questions we defined five categories of increase suicidality: “no suicidality” controls, “thought life not worth living,” “contemplated self-harm or suicide,” “acts of deliberate self-harm not including attempted suicide,” and “attempted suicide.” Controls were required to have answered each question with the least severe option *(e.g*., “no” in response to “Many people have thoughts that life is not worth living. Have you felt that way?”). If multiple suicidality questions were endorsed, the UKB participant was assigned to the more severe suicidality group. In accordance with our interest in understanding the environmental impact on thoughts and behaviors leading up to death by suicide, samples with death by suicide as their cause of death (ICD10 codes X60-X84) and those who did not answer any of the suicidality questions were removed from the analysis.

### UKB Environments

To study the effects of traumatic events, responses to trauma, and diagnosis of posttraumatic stress disorder (PTSD) we included 26 environments from the UKB mental health questionnaire.^85^

#### Traumatic events and posttraumatic stress

A total of 21 items related to exposure to traumatic events category of the UKB mental health questionnaire describing three phenotype classes: (1) information regarding exposure to traumatic events, (2) behavioral and emotional responses to trauma, and (3) information regarding social support. In addition to the social support domains included in the traumatic events category, we included the UKB sociodemographic information for *Townsend deprivation index* (UKB Field ID: 189) and *average total household income before tax* (UKB Field ID: 738). The Townsend deprivation index describes material deprivation within a population as a single continuous measure of unemployment, automobile non-ownership, home non-ownership, and household overcrowding. Prior studies from our group have shown that these are strong mediators of relationships between PTSD and other mental health, anthropometric, and inflammatory phenotypes.^56, 57, 86^ Finally, we included one phenotype from the depression category of the mental health questionnaire: *Over the last two weeks, how often have you had trouble concentrating on things, such as reading the newspaper of watching television* (UKB Field ID: 20508).

#### PTSD symptom severity count and diagnosis

Information to screen for PTSD was available for >150,000 UKB participants. We used six items from the online mental health questionnaire to derive PTSD diagnoses in the UKB sample including questions related to avoidance of activities, repeated disturbing thoughts, feeling upset, feeling distant from others, feeling irritable, recent trouble concentrating (UKB Field IDs 20494-20498 and 20508)^85^ The first five items were scored on a five-point scale according to the amount of concern caused by that item and ranged from 1=“not at all” to 5=“extremely”. The final item about concentration was scored on a four-point scale describing the frequency of difficult associated with that item (1=“not at all” to 4=“nearly every day”). The responses to these questions were summed to produce a single continuous score ranging from 3-29. Those UKB participants with PCL-6 scores ≥13 were defined as probably PTSD cases.^85^

#### Imputation

Environments were imputed using the MICE (Multivariate Imputation via Chained Equations)^17^ package in R using ten iterations of 5 multiple imputations. Briefly, every missing value is imputed with placeholders followed by sequential resetting of each missing variable to “missing.” Then the observed values for missing environments were regressed on other environments in the imputation model (*e.g*., the missing environment is the dependent variable). Missing values in the imputed environment are then replaced with predicted from the regression model. This process was performed to fill all missing environmental variables. To evaluate imputation reliability, we compared the relative differences in standard error estimates for the distribution of original phenotypes, each imputed phenotype distribution, and the final phenotype distribution. Imputation accuracies were evaluated by comparing the relative change in standard error (dSE) between original unimputed and imputed data. All Yale-Penn environments exhibited a reduction on standard error (range = −0.04% to −48.5%) indicating that they were reliably imputed (Table S1 and S2).^16^

### Heritability and Genetic Correlation

Observed-scale *h*^2^ was calculated for munged GWAS summary statistics using the Linkage Disequilibrium Score Regression (LDSC)^18^ method with the 1000 Genomes Project European population as a linkage disequilibrium reference. Genetic correlation between phenotypes also was calculated with LDSC for all phenotypes pairs in which each trait has an *h*^2^ z-score ≥ 4.

### Gene-by-environment genome-wide interaction analyses (GEWIS)

GEWIS was performed using the recently developed StructLMM,^15^ a linear mixed-model approach to efficiently detect interactions between loci and one or more environments. Here we used the StructLMM approach to evaluate whether traumatic events, response to trauma, PTSD, or socioeconomic status individually and collectively interact with respect to risk for suicidality. In the StructLMM framework, loci with significant additive or interactive relationships with environments were analyzed *post hoc*.

First, to estimate the model importance we calculated marginal log model likelihoods (log(Bayes factor *(BF))* between the full model and the reduced model with environments removed to identify which environments are most relevant for the detected locus interaction effects. To prioritize a putative causal set of environments, we performed a greedy backwards elimination procedure prioritizing sequential removal of the strongest individual mediating environments until the change in log marginal likelihood (Δlog(*BF*)) < 1.^15^

Second, we performed allele effect prediction with the StructLMM `OptimalRho` function whereby 50% of the UKB participants were used to train an allele effect prediction and the remaining 50% were used as the test sample. Effect predictions are based on the best linear unbiased predictor (BLUP)^87^ accounting for the different relative importance of each environment. Specific details of the statistics and methods employed by StructLMM have been described previously^15^ StructLMM GEWIS of suicidality was performed using age, genotyping batch, and 10 principal components of ancestry as covariates. For the full sample combining males and females, we also included sex as a covariate.

### Functional Annotation of GEWIS

Association results from GEWIS were mapped to genes using MAGMA^29^ implemented in FUMA^30, 31^ based on positional mapping of each variant to genes in a 2kb window upstream and downstream of the variant *(i.e*., 4kb window size).^30, 31^ With this tool we evaluated enrichment of gene-ontology, tissue-transcriptomic, and cell-type specific transcriptomic profiles. Tissue transcriptomic profile enrichments were analyzed using GTEx v8. Cell-type transcriptomic profile enrichments were evaluated with the following human brain annotations: PsychENCODE_Developmental,^88^ PsychENCODE_Adult,^88^ Allen_Human_LGN_level1,^60^ Allen_Human_LGN_level2,^60^ Allen_Human_MTG_level1,^60^ Allen_Human_MTG_level2,^60^ DroNc_Human_Hippocampus,^89^ GSE104276_Human_Prefrontal_cortex_all_ages,^90^ GSE104276_Human_Prefrontal_cortex_per_ages,^90^ GSE67835_Human_Cortex,^91^ GSE67835 Human Cortex woFetal,^91^ Linnarson GSE101601 Human Temporal cortex,^92^ Linnarson_GSE76381_Human_Midbrain. Enrichments and gene-based tests were adjusted for multiple testing using a false discovery rate of 5%.

Further annotation of sex-specific gene sets was performed using the ShinyGO platform (v0.61)^32^

### GWAS versus GEWIS Comparison

First, we evaluated the three LD-independent GWS SNPs from Strawbridge, *et al*.^7^ in our association and interactive tests using the same set of environments and covarying for age, sex, 10 PCs, and genotype batch. We then performed `bf` and `OptimalRho` analyses to characterize the GxE effects at these loci. Second, we looked up the GEWIS loci from association and interaction tests in our discovery cohort in the summary statistics from Strawbridge, *et al*.^7^

### Replication

We replicated the GxE effects of genomic risk regions discovered from GEWIS association and interaction tests in our discovery cohort using the Yale-Penn cohort. Yale-Penn participants were collected from five study sites in the eastern United States for studies of the genetics of substance dependence and other psychiatric and behavioral traits using the Semi-Structured Assessment for Drug Dependence and Alcoholism (SSADDA).^94, 95^ Participants provided written informed consent through a protocol approved by the institutional review board at each participating site (Yale Human Research Protection Program (protocols 9809010515, 0102012183, and 9010005841), University of Pennsylvania Institutional Review Board, University of Connecticut Health Center Institutional Review Board, Medical University of South Carolina Institutional Review Board for Human Research, and the McLean Hospital Institutional Review Board). To replicate our GEWIS findings, we focused on European ancestry Yale-Penn subjects with unambiguously defined sex assignments (*N*=3,099, 37.5% female).

We mapped the four questionnaire items used to derive the suicidality phenotype in UKB to four comparable phenotypes in Yale-Penn (all answered in a binary yes/no manner): *“Did you have thoughts of dying, or taking your own life, or wishing you were dead?” “Have you ever thought about killing yourself?” “Did you ever hurt yourself on purpose (e.g., by cutting or burning)?”* and *“Have you ever tried to kill yourself?”* Among the total 3,099 Yale-Penn replication samples, 1,485 individuals were coded as *“no suicidality”* controls, 225 as *“thought life not worth living,”* 751 as *“contemplated self-harm or suicide,”* 245 as *“deliberate self-harm,”* and 393 as *“attempted suicide.”*

Ten environments from Yale-Penn were included in the replication GEWIS, which represent the same domains tested in the discovery cohort: military exposure, posttraumatic stress, socioeconomic status, and social support. These phenotypes are described in Table S24. Imputed environments from Yale-Penn showed no evidence of increased dSE suggesting that the data were reliably imputed (−48.3≤dSE≤-5.19).^16, 87^

Replication was performed for all five GEWIS-discovered genomic risk regions in the UKB (Tables S5-S9). Where Yale-Penn did not contain suitable information for the genomic risk region, we performed GEWIS for the first 75 SNPs up and downstream of the region. For example, the chromosome 9 genomic risk region tests 150 SNPs from chr9:137112902-chr9:13749377 (Table S9).

## Data Availability

Summary level data used in the project are made available for download on the UKB data access portal (https://biobank.ctsu.ox.ac.uk/crystal/browse.cgi?id=-2&cd=search) or from their referenced publications. Individual level data are available from UKB to bona fide researchers through the UKB Access Management System.

https://biobank.ctsu.ox.ac.uk/crystal/browse.cgi?id=-2&cd=searc

http://researchdata.gla.ac.uk/930/

## Acknowledgements

This study was supported by the National Center for PTSD of the U.S. Department of Veterans Affairs, the American Foundation for Suicide Prevention (YIG-1-109-16), the National Institutes of Health (F32 MH122058, R21 DC018098, R21 DA047527, R01 DA12690), and the U.S. Department of Veterans Affairs Office of Academic Affiliations Advanced Fellowship Program in Mental Illness Research and Treatment and the VISN 4 Mental Illness Research, Education and Clinical Center. The content is solely the responsibility of the authors and do not necessarily represent the views of the funding agencies. This research has been conducted using the UK Biobank Resource (application reference no. 58146).

## Author Contributions

FRW and RP conceived the study. FRW, GAP, DFL, YZN, CO, and CT, contributed to phenotype mapping across cohorts. FRW performed all study analyses with support from KA. FRW drafted the original manuscript. FRW GAP, DFL, YZN, CO, CT, KA, FDA, DST, AG, JHK, CGA, MBS, HRK, JG, and RP critically evaluated the manuscript and interpretation of findings. RP received funding support.

